# Mortality Associated With Intubation and Mechanical Ventilation in Patients with COVID-19

**DOI:** 10.1101/2020.08.13.20174524

**Authors:** Patrick W. Zimmerman, Stephanie J. Stroever, Timothy Burton, Karri E. Hester, Minha Kim, Ryan T. Fahy, Kimberly A. Corbitt, Joann R. Petrini, Jeffrey Nicastro

## Abstract

**Objective:** The need for critical care, hemodynamic support, renal replacement therapy, and mechanical ventilation have emerged as key features of the SARS-nCoV-2 (COVID-19) pandemic. The primary aim of this study was to determine the in-hospital mortality rate of mechanically ventilated patients. We also sought to determine the risk of in-hospital mortality by age, gender, race, ethnicity, and body mass index.

**Methods:** We performed a retrospective cohort study to determine the mortality rate among inpatient adults with COVID-19 on mechanical ventilation in the Nuvance Health system between March 1, 2020 and July 17, 2020. Patients were included if they were 18 years or older, had a laboratory confirmed COVID-19 diagnosis, were admitted to hospitals within the Nuvance Health network (7 hospitals), and were on mechanical ventilation at any time during their inpatient stay.

**Results:** Overall mortality in our cohort of 304 patients was 53.3%. Multivariable logistic regression including age, gender, race, ethnicity, and BMI demonstrated patients over 71 years old had greater risk of mortality compared to patients ages 61-70, and females had half the risk compared to males. There was no significant difference in risk of mortality given race, ethnicity, or BMI.

**Conclusions:** In adult patients with confirmed COVID-19 infection requiring mechanical ventilation and intensive care, advanced age (>71 years old) and male gender are associated with increased risk of mortality. This information contributes to a collective body of evidence to support ongoing planning and decision-making among clinicians and for directed infection prevention programming.

## Introduction

The novel coronavirus causing COVID-19 (SARS-CoV-2) was first detected in Wuhan, China in December 2019. Since that time, the disease has spread worldwide with more than 14 million confirmed cases and over 605,000 deaths [1]. New York City was one of the first epicenters of disease in the United States [2]. As a result, neighboring regions, including western Connecticut and eastern New York, experienced high volumes of COVID-19 positive patients. As of July 17, 2020, Connecticut reported 45,782 positive cases and 4,263 deaths, while Putnam and Dutchess Counties in eastern New York reported 1,305 and 4,150 positive cases respectively [3,4]. The Nuvance Health network, a healthcare system situated in this region, cared for more than 1,500 COVID-19 positive patients between March and July 2020.

COVID-19 can cause asymptomatic, mild, and severe disease. Acute respiratory failure is the hallmark of severe disease, though there are also extensive reports of acute kidney injury, shock, organ dysfunction, acute cardiac injury, and hypercoagulation leading to arterio-occlusive events [5-10]. Critical care has proven to be essential in the COVID-19 crisis and many critical patients require invasive mechanical ventilation for respiratory support [2,6,10-14]. Singer et al. reported approximately 19% of patients treated in a large, academic medical center in New York required mechanical ventilation [2]. Large facilities in Georgia and Detroit, Michigan also reported high rates of mechanical ventilation [6,13]. Regrettably, mortality rates among critically ill patients have been reported between 15.4 and 61.5% worldwide [6,8,12,14,15].

Though there are many comprehensive manuscripts that describe the characteristics of COVID-19 patients, there is still an urgent need for data that estimates the risk of mortality in different samples of the population. Generalizability is difficult with limited data. Thus, it is important for hospitals and healthcare systems treating large numbers of patients to provide details to further the collective knowledge of the disease. The primary objective of this study was to provide in-hospital mortality rates of patients who required mechanical ventilation during treatment for COVID-19. We also sought to estimate the risk of mortality across demographic characteristics of our patients, including age, race and ethnicity, gender, and body mass index (BMI).

## Materials and Methods

### Study Population, Setting, and Data Acquisition

We conducted a retrospective cohort study to determine the in-hospital mortality rate of COVID-19 positive patients on mechanical ventilation in the Nuvance Health system. Nuvance Health includes seven hospitals spanning two states and five counties. It serves a diverse population of approximately 1.5 million patients in New York and Connecticut.

We included patients in our study if they were 18 years or older with RT-PCR confirmed COVID-19, admitted to any hospital in the Nuvance Health system, and were on mechanical ventilation at any time during their inpatient stay between March 3 and July 17, 2020. The primary drivers for intubation and initiation of mechanical ventilation were moderate to severe acute respiratory failure with hypoxia, hypoxemia, or hypercapnea unresponsive to high-flow oxygen therapy (15L by non-rebreather mask). We excluded patients that were pregnant, on hospice or comfort care only, or expired before testing for COVID-19.

We selected patients via the Nuvance Health electronic medical record system and billing department based on the COVID-19 ICD-10 code (U07.1) and positive RT-PCR results. Investigators gathered and compiled the appropriate data using the Research Electronic Data Capture application (REDCap) [16,17].

### Variables

The primary outcome for this study was in-hospital mortality defined as expiration prior to the date of discharge. Covariates included age, race and ethnicity, gender, and BMI. We measured age as a continuous variable, though used a categorical variable in increments of 10 years for multivariable modeling. We defined gender as male or female and race as White, Black or African American, Asian and Native Hawaiian/Pacific Islander, and other. The race category of “other” was created combining patients who were described in the hospital electronic medical record as “other,” “did not identify” or were Indian (from India). We defined ethnicity as non-Hispanic or Hispanic, and categorized patients who did not identify as “unknown.” Lastly, we included BMI as a continuous variable for descriptive statistics. However, we categorized BMI for multivariable analyses according to the Centers for Disease Control and Prevention definitions for overweight and obesity (Table 1) [18].

**Table 1.**
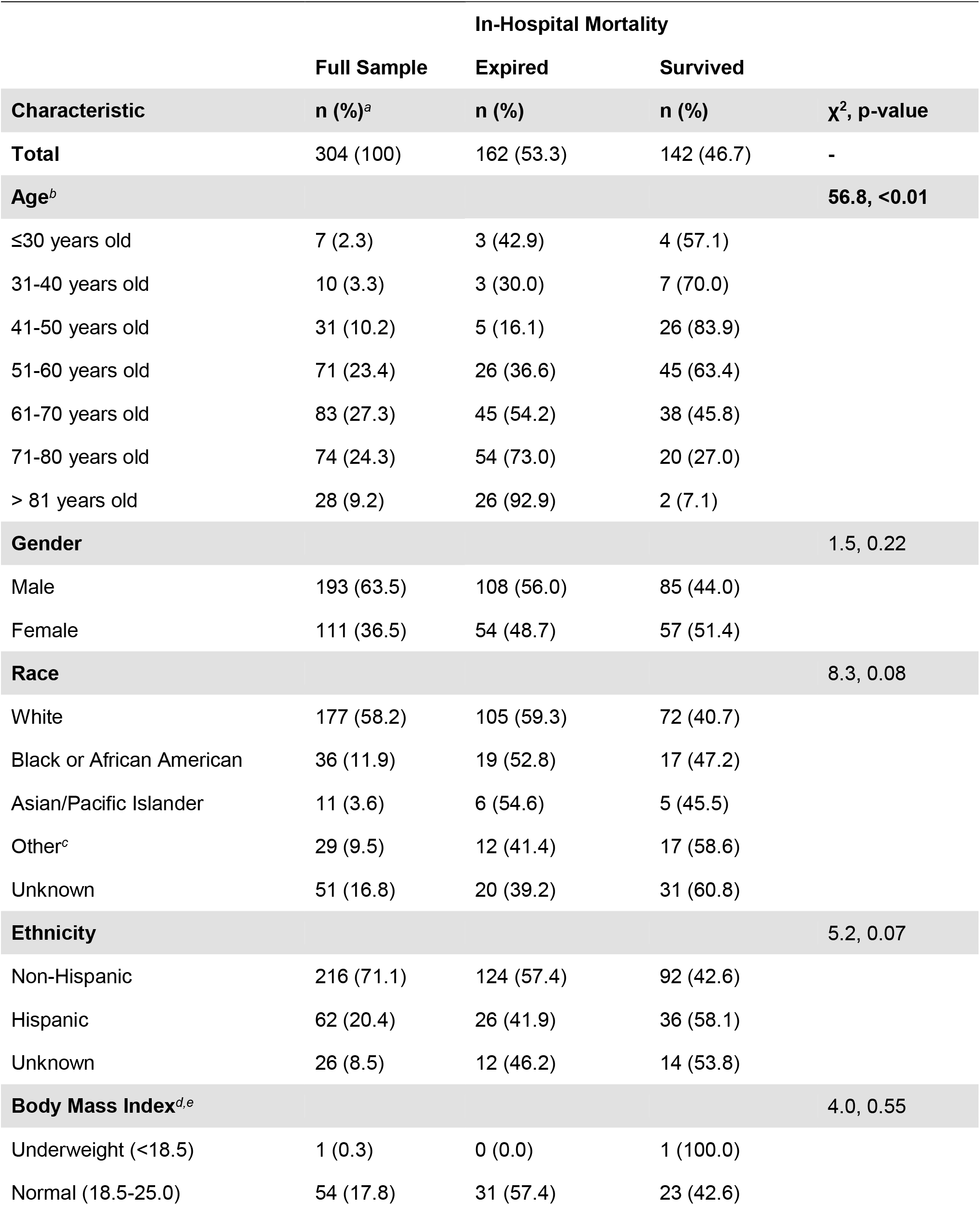
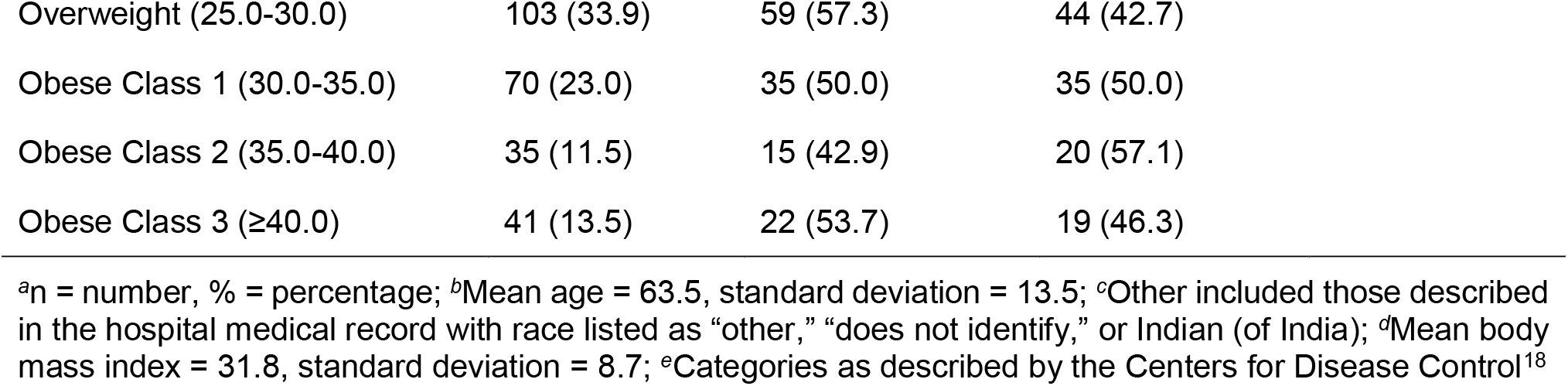
Descriptive statistics and univariate differences in mortality among COVID-19 positive patients that required mechanical ventilation at Nuvance Health, March 3-July 17, 2020 (N = 304).

| Characteristic | In-Hospital Mortality | | | $\chi^2$, p-value |
| --- | --- | --- | --- | --- |
|  | Full Sample<br>n (%)<sup>a</sup> | Expired<br>n (%) | Survived<br>n (%) |  |
| **Total** | 304 (100) | 162 (53.3) | 142 (46.7) | - |
| **Age<sup>b</sup>** |  |  |  | **56.8, <0.01** |
| ≤30 years old | 7 (2.3) | 3 (42.9) | 4 (57.1) |  |
| 31-40 years old | 10 (3.3) | 3 (30.0) | 7 (70.0) |  |
| 41-50 years old | 31 (10.2) | 5 (16.1) | 26 (83.9) |  |
| 51-60 years old | 71 (23.4) | 26 (36.6) | 45 (63.4) |  |
| 61-70 years old | 83 (27.3) | 45 (54.2) | 38 (45.8) |  |
| 71-80 years old | 74 (24.3) | 54 (73.0) | 20 (27.0) |  |
| > 81 years old | 28 (9.2) | 26 (92.9) | 2 (7.1) |  |
| **Gender** |  |  |  | **1.5, 0.22** |
| Male | 193 (63.5) | 108 (56.0) | 85 (44.0) |  |
| Female | 111 (36.5) | 54 (48.7) | 57 (51.4) |  |
| **Race** |  |  |  | **8.3, 0.08** |
| White | 177 (58.2) | 105 (59.3) | 72 (40.7) |  |
| Black or African American | 36 (11.9) | 19 (52.8) | 17 (47.2) |  |
| Asian/Pacific Islander | 11 (3.6) | 6 (54.6) | 5 (45.5) |  |
| Other<sup>c</sup> | 29 (9.5) | 12 (41.4) | 17 (58.6) |  |
| Unknown | 51 (16.8) | 20 (39.2) | 31 (60.8) |  |
| **Ethnicity** |  |  |  | **5.2, 0.07** |
| Non-Hispanic | 216 (71.1) | 124 (57.4) | 92 (42.6) |  |
| Hispanic | 62 (20.4) | 26 (41.9) | 36 (58.1) |  |
| Unknown | 26 (8.5) | 12 (46.2) | 14 (53.8) |  |
| **Body Mass Index<sup>d,e</sup>** |  |  |  | **4.0, 0.55** |
| Underweight (<18.5) | 1 (0.3) | 0 (0.0) | 1 (100.0) |  |
| Normal (18.5-25.0) | 54 (17.8) | 31 (57.4) | 23 (42.6) |  |

|  |  |  |  |
| --- | --- | --- | --- |
| Overweight (25.0-30.0) | 103 (33.9) | 59 (57.3) | 44 (42.7) |
| Obese Class 1 (30.0-35.0) | 70 (23.0) | 35 (50.0) | 35 (50.0) |
| Obese Class 2 (35.0-40.0) | 35 (11.5) | 15 (42.9) | 20 (57.1) |
| Obese Class 3 ($\geq 40.0$) | 41 (13.5) | 22 (53.7) | 19 (46.3) |

<sup>a</sup>n = number, % = percentage; <sup>b</sup>Mean age = 63.5, standard deviation = 13.5; <sup>c</sup>Other included those described in the hospital medical record with race listed as “other,” “does not identify,” or Indian (of India); <sup>d</sup>Mean body mass index = 31.8, standard deviation = 8.7; <sup>e</sup>Categories as described by the Centers for Disease Control<sup>18</sup>

**Table 2.** Risk differences in mortality among COVID-19 positive patients that required mechanical ventilation at Nuvance Health, March 3-July 17, 2020 (N = 304).

| Variable | Adjusted Odds Ratio<sup>a</sup> | 95% Confidence Interval | P-value |
| --- | --- | --- | --- |
| **Age** |  |  |  |
| <30 years old | 0.54 | 0.10, 2.81 | 0.46 |
| 31-40 years old | 0.41 | 0.09, 1.82 | 0.24 |
| 41-50 years old | 0.17 | 0.06, 0.52 | **<0.01** |
| 51-60 years old | 0.45 | 0.23, 0.89 | **0.02** |
| 61-70 years old | Reference | - | - |
| 71-80 years old | 2.31 | 1.14, 4.68 | **0.02** |
| > 81 years old | 13.9 | 2.97, 64.87 | **<0.01** |
| **Gender** |  |  |  |
| Male | Reference | - | - |
| Female | 0.47 | 0.27, 0.85 | **0.01** |
| **Race** |  |  |  |
| White | Reference | - | - |
| Black or African American | 1.48 | 0.61, 3.61 | 0.39 |
| Asian or Pacific Islander | 0.89 | 0.28, 2.80 | 0.84 |
| Other | 1.45 | 0.30, 6.90 | 0.64 |
| Unknown | 1.01 | 0.33, 3.13 | 0.99 |
| **Ethnicity** |  |  |  |
| Non-Hispanic | Reference | - | - |
| Hispanic | 1.19 | 0.45, 3.12 | 0.72 |
| Unknown | 1.09 | 0.37, 3.18 | 0.87 |
| **Body Mass Index** |  |  |  |
| Underweight (<18.5) | Dropped<sup>b</sup> | - | - |
| Normal (18.5-25.0) | Reference | - | - |
| Overweight (25.0-30.0) | 1.22 | 0.56, 2.64 | 0.62 |
| Obese Class 1 (30.0-35.0) | 1.01 | 0.44, 2.35 | 0.97 |
| Obese Class 2 (35.0-40.0) | 0.97 | 0.35, 2.64 | 0.95 |
| Obese Class 3 (≥40.0) | 1.31 | 0.50, 3.42 | 0.58 |

<sup>a</sup>Adjusted odds ratios from multivariable logistic regression that included age, gender, race, ethnicity, and body mass index; <sup>b</sup>Observation for underweight body mass index dropped because it predicted failure perfectly in the model (n = 1)

### Statistical Analysis

We conducted statistical analyses using StataSE version 16 [19]. There was no missing data in the sample requiring correction. We calculated the crude overall mortality rate for all patients in the cohort and stratified mortality by age, race, ethnicity, gender, and BMI. We also performed univariate chi-square analyses to determine group differences in mortality for each covariate. Lastly, we conducted multivariable logistic regression to define the risk differences of mortality for each covariate while controlling for the others. We used the most prevalent age group (61-70 years old) as the reference group for age and normal weight as the reference group for BMI. We verified all assumptions of logistic regression and established an alpha of 0.05 *a priori* for statistical significance.

## Results

### Descriptive statistics

There were 1,683 patients admitted to Nuvance Health hospitals between March 3 and July 17, 2020. A total of 304 (18.1%) adults with laboratory confirmed COVID-19 required mechanical ventilation during their inpatient stays (Table 1). The majority of patients were male (63.5%), non-Hispanic (71.1%) and White (58.2%). The mean age of the sample was 63.5 (standard deviation = 13.5) with 60.8% of the sample over 60 years. Additionally, the mean BMI was 31.8 (standard deviation = 8.7).

### Univariate group differences

The overall mortality in our cohort was 53.3%. There was a statistically significant difference in mortality across age groups (p < 0.01). The highest mortality rates were among patients over age 71 with more than 70% mortality in each group. The lowest mortality rate was among patients 41 to 50 years old (16.1%). COVID-19 mortality rates also differed by race with rates being highest for White and African American patients (59.3% and 52.8%, respectively) and the lowest rate for those categorized as “unknown” (39.2%). There were no statistically significant differences across race, ethnicity, or gender on univariate analysis. Lastly, there was not a statistically significant difference in mortality across categories of overweight and obesity (X^2^ = 4.0, 0.55).

### Multivariable logistic regression

The risk of death among mechanically ventilated patients ages 71-80 was 2.31 times higher (95% CI = 1.14, 4.68, p = 0.02) than patients ages 61-70 after controlling for gender, race, ethnicity, and BMI. Additionally, patients over age 81 were at a much higher risk of mortality (aOR = 13.9, 95% CI = 2.97, 64.87, p <0.01) than patients ages 61-70, though the wide confidence interval suggests challenges with model estimation in this group. Unlike the univariate analysis, females were at significantly lower risk of mortality than males (aOR = 0.47, 95% CI = 0.27, 0.85, p = 0.01) after controlling for other variables in the model. The difference in statistical significance between the univariate and multivariable models is likely the result of negative confounding, in which failing to control for age in the univariate model minimized the difference between males and females. Controlling for age permitted a more accurate estimation of the risk differences between genders. Lastly, we did not find significant differences in the risk of mortality across race after controlling for other demographic variables.

## Discussion

The purpose of this retrospective cohort study was to describe the rate and risk of mortality in 279 critically ill adult inpatients who required mechanical ventilation for laboratory confirmed COVID-19 infection. Our overall mortality was high at 52%, though it was lower than that of early reports from China and did not reach the heights reported in Seattle [5,7,9,20,21]. Additionally, our rate was similar to reports from New York City [2]. The generalizability of the data was unclear from the early U.S. (Seattle) data, as well as that of Italy, China, and others. The consistency of data from our region and New York City despite differing demographics is reassuring in terms of generalizability.

The majority of deaths occurred in patients over age 60 (60.8%). There was a clear increase in the risk of mortality among older patients which is consistent with current COVID-19 literature for adults [5-9, 12, 13, 15, 21,22]. Suleyman et al. reported the odds of death in patients over 60 were 5.3 times that of patients less than 60 years (p < 0.001) [6]. Similarly, Cummings et al. reported an adjusted hazard ratio of 1.31 for every 10-year increase in age (95% CI = 1.09, 1.57), while Zangrillo et al. reported an odds ratio of 1.12 (p = 0.004) [12,15]. These findings are not surprising, as older adults tend to have less physiologic reserve and more co-morbidities. There is a need for further research to identify the mechanism of COVID-19 that places older patients at greater risk.

Surprisingly, we also found 42.9% mortality among patients 30 years or younger. This is different from prior work that demonstrated mortality of 0% among critically ill patients under 30, and 8.8% in patients 25-45 years old [2,8]. Though the sample size in this sub-group was small (n = 6), clinicians should be aware of this finding given the increasing number of COVID-19 cases in younger adults. Further research into the mechanism of death in this age group is also warranted, as it is likely different from older adults.

In our cohort, women had lower mortality than men. This is consistent with prior literature, though the reason behind the finding is poorly understood. Investigators are starting to assess biological differences between males and females, particularly the expression of angiotensin-converting enzyme 2 (ACE2) and the immune response [23-25]. Additional research is needed to further understand this disparity in mortality risk among different sexes.

We did not find significant differences in mortality rates or risk of death across race and ethnicity. Our findings are similar to other hospital-based studies in the United States [6,26]. Population-based studies do report disparities in outcomes among different races. Sehra et al. used state-level data from New York to assess risk differences in mortality by race, obesity, age, and other state-level factors [27]. They found a significantly increased risk of mortality due to COVID-19 among African Americans. Holmes, Jr. et al. also demonstrated differences in mortality using state-level data [28]. The differences between hospital and state-level data suggest that differences in access to care or social determinants of health may play a role in this pandemic. Tai et al. suggest clinicians work with communities to reduce barriers to care among racial minorities [29].

Lastly, our study did not find a significant difference in the rate or risk of mortality among obese patients. Our findings are consistent with other reports from New York that did not find obesity to be an independent risk factor for mortality, though reports from reviews suggest obesity does play a role in mortality [12,30,31]. Importantly, our study only included critically ill patients. It is possible our study was underpowered to find small differences in risk between obese and normal weight patients among the critically ill. These findings may be different in a larger sample with patients across severity spectrum. There may also be important moderators of the association with mortality, such as diabetes or hypertension that were not included in this study. Further research is needed to clarify the impact of obesity in patients with COVID-19.

## Limitations

Our study has several limitations. First, our sample is relatively homogenous. More than half of patients in our sample were non-Hispanic and White. Of note, our sample included a large proportion of patients who did not identify as a specific race or ethnicity. We also had numerous patients listed as “other” in the medical record. Our results may reflect a misclassification bias that influenced the result towards the null. Additionally, our study did not include other risk factors of mortality for COVID-19. There may be important clinical factors (i.e., comorbidities and secondary sequelae) that play an unmeasured role in the reported risk and rate of mortality. However, our data is useful in the context of observational meta-analyses and in communities with similar demographics.

## Conclusions

The COVID-19 pandemic has caused widespread morbidity and mortality in the United States. As the nation continues to combat the pandemic with public health programs and legislative intervention, clinicians continue to battle the disease. Though the evidence base is much more robust than at the onset of disease, there is still much to learn about the management of the disease in high risk groups. The information from this study can provide much needed support in clinical decision-making, resource allocation, and directed infection prevention programs.

## Declarations

### Ethics Approval

This study was performed in line with the principles of the Declaration of Helsinki. Approval was granted by the Biomedical Research Alliance of New York Institutional Review Board (IRB # 20-12-108-337(c20)) and the Vassar Brothers Medical Center Institutional Review Board (DCR# 2020-2).

### Consent to participate

The study was approved as exempt under category #4(iii) as detailed in 45 CFR 46.104(d) as secondary research for which consent is not required.

## Data Availability

Data is available upon request to the corresponding author.

## Author Contributions

P. Zimmerman, J. Nicastro, and S. Stroever were involved in the conception and design of the research study. P. Zimmerman, T. Burton, K. Hester, M. Kim, R. Fahy, and K. Corbitt were involved in data acquisition and revision of the manuscript. S. Stroever was involved in data analysis and interpretation, drafting, and revision of the manuscript. J. Petrini and J. Nicastro were involved in critical revision of the manuscript.

## Acknowledgments

Jonathan Serino, Department of Research and Innovation at Nuvance Health; Beth Falder, Quality Analytics at Nuvance Health; Ray Pankuch and Information Technology colleagues at Nuvance Health; Laura Morales, MD, Helene Rached, MD, and William Wing, MD of the Department of Surgery, Vassar Brothers Medical Center, Nuvance Health

## Take Home Message

Approximately half of the patients on mechanical ventilation due to COVID-19 died from the disease, with greater risk among males and older patients. Obesity was not a significant risk factor for mortality in this specific cohort.

